# Gastric dysfunction in patients with chronic nausea and vomiting syndromes defined by a novel non-invasive gastric mapping device

**DOI:** 10.1101/2022.02.07.22270514

**Authors:** Armen A. Gharibans, Stefan Calder, Chris Varghese, Stephen Waite, Gabriel Schamberg, Charlotte Daker, Peng Du, Saeed Alighaleh, Daniel Carson, Jonathan Woodhead, Gianrico Farrugia, John A. Windsor, Christopher N. Andrews, Greg O’Grady

## Abstract

**Importance:** Chronic nausea and vomiting syndromes (NVS) are prevalent and debilitating disorders. Putative mechanisms include gastric neuromuscular disease and dysregulation of brain-gut interaction, but clinical tests for objectively defining gastric motor function are lacking.

**Objective:** A novel medical device enabling non-invasive body surface gastric mapping (BSGM) was developed and applied to evaluate NVS pathophysiology.

**Design:** A case-control study where BSGM was performed in NVS patients and matched controls using Gastric Alimetry (Alimetry, New Zealand), comprising a conformable high-resolution array (8×8 electrodes; 20 mm inter-electrode spacing), wearable Reader, and validated symptom logging App. Continuous measurement encompassed a fasting baseline (30 min), 482 kCal meal (10 min), and 4-hr post-prandial recording.

**Setting:** Multicenter study in Auckland, New Zealand and Calgary, Canada.

**Participants:** 43 NVS patients (gastroparesis and Rome IV chronic NVS) and 43 matched controls.

**Main outcomes and measures:** Symptom severity and quality of life were measured using Patient Assessment of Upper Gastrointestinal Disorders-Symptom Severity Index (PAGI-SYM), Gastroparesis Cardinal Symptom Index (GCSI), and Patient Assessment of Upper Gastrointestinal Disorders-Quality of Life (PAGI-QOL) instruments. Health psychology metrics included the State Trait Anxiety Inventory Short Form (STAI-SF) and Patient Health Questionnaire-2 (PHQ-2) questionnaires. Spectral analyses including frequency, amplitude, and fed-fasting power ratio. Spatial biomarker analyses included spatial frequency stability and average spatial covariance.

**Results:** Meal responses were impaired in NVS, with reduced amplitudes compared to controls (median 23.3 vs 38.0 µV, p<0.001), impaired fed-fasting power-ratios (1.1 vs 1.6, p=0.02), and disorganized slow-waves (spatial frequency stability 13.6 vs 49.5; p<0.001). However, two distinct NVS subgroups were evident with indistinguishable symptoms (all p>0.05). A majority (62%) had normal BSGM studies (all biomarkers non-significant vs controls) with increased psychological comorbidities (43.5% vs 7.7%; p=0.03) and anxiety scores (median 16.5 vs 13.0; p=0.035). A smaller subgroup (31%) had markedly abnormal BSGM, with test biomarkers correlating with symptoms (nausea, pain, excessive fullness, early satiety, bloating; all r>0.35, p<0.05).

**Conclusions and Relevance:** NVS patients share overlapping symptoms, but comprise distinct underlying phenotypes as revealed by a novel BSGM device. These phenotypes correlate with symptoms, which should inform clinical management and allocations into therapeutic trials.

**Key Points:** *Question:* How does body surface gastric mapping, a novel non-invasive medical device for evaluating gastric motility, aid assessment of patients with chronic nausea and vomiting.

*Findings:* Two subgroups were revealed in chronic nausea and vomiting syndromes, which could not be differentiated by symptoms alone. Where body surface gastric mapping was normal, symptoms correlated with psychological comorbidities, and where body surface gastric mapping was abnormal, symptoms correlated with gastric electrophysiology metrics.

*Meaning:* Distinct phenotypes revealed by body surface gastric mapping correlate with symptoms, which should inform targeted clinical management and allocations into therapeutic trials.

## Introduction

Gastroparesis and chronic nausea and vomiting syndrome (CNVS; defined by Rome IV Criteria) are debilitating disorders with a combined global prevalence of >1%.^1–3^ These disorders overlap, sharing similar epidemiology, symptomatology, and quality of life impacts, and impart a growing healthcare burden.^3–7^ Although differentiated by the presence or absence of delayed gastric emptying, this distinction is controversial because gastric emptying is unstable over time, correlates weakly with symptoms, and may not reflect the primary disease mechanism.^8–12^ The two disorders may therefore be considered together under the umbrella term nausea and vomiting syndromes (NVS).^13^

The heterogeneous and unclear pathophysiology of NVS contributes to diagnostic uncertainty and impedes therapeutic progress. Contributing mechanisms may include gastric neuromuscular dysfunction, dysregulation of gut-brain interaction, and pylorospasm, which could represent distinct or overlapping phenotypes.^7,14,15^ Previous investigations have identified gastric neuromuscular pathologies in subsets of patients with both gastroparesis and CNVS, suggesting they may lie on the same disease spectrum.^7,11,16–18^ A dominant feature has been interstitial cell of Cajal (ICC) depletion, associated with changes in proinflammatory macrophages,^11,16,17,19^ which has been linked to aberrant gastric electrical patterns recorded using high-resolution (HR) electrical mapping.^16,17,20,21^

Electrogastrography (EGG) has previously been the only clinical method for evaluating gastric myoelectrical function. EGG has demonstrated consistent abnormalities in NVS,^13^ however multiple limitations restricted clinical adoption, including noise susceptibility, low resolution in the face of gastric anatomical variability, inability to evaluate spatial patterns, and low specificity for disease subtypes.^22,23^ Body surface gastric mapping (BSGM; also HR-EGG) has recently emerged as a new approach, employing dense fields of electrodes to reliably assess gastric activity at high spatial resolution.^22^ Recent studies have shown that BSGM biomarkers achieve improved sensitivity and superior symptom correlations compared to EGG and gastric emptying testing.^24,25^ BSGM has been comprehensively validated, and a scalable platform introduced, now enabling clinical translation.^26,27^

The aim of this study is to present a novel medical device and clinical procedure for non-invasive BSGM (Gastric Alimetry^®^; Alimetry, New Zealand), and the results of its first application in patients with NVS.

## Methods

This was a case-control study conducted in Auckland, New Zealand, and Calgary, Canada. Ethical approvals were given by The Auckland Health Research Ethics Committee (AHREC; AH1130) and Conjoint Health Research Ethics Board (CHREB; REB19-1925). All patients provided informed consent. The study is reported in accordance with the STROBE Statement.^28^

### Participants

Patients aged ≥18 years with a clinical diagnosis of CNVS (Rome IV Criteria^29^) or gastroparesis were eligible. Patients were evaluated and recruited by gastroenterologists, and all were tested to rule out alternative causes for symptoms. Testing included endoscopy and scintigraphic gastric emptying. Exclusion criteria were metabolic, neurogenic, or endocrine disorders known to cause gastric dysmotility other than diabetes (e.g., scleroderma, multiple sclerosis, hyperthyroidism), an active gastrointestinal infection (including H. pylori), inflammatory bowel disease, previous gastric or esophageal surgery, history of gastrointestinal malignancy, or current pregnancy. Patients with cyclical vomiting syndrome or cannabinoid hyperemesis were excluded. Patients were matched to a database of controls (115 subjects ≥18 years recruited by local advertisement during 2021) in a 1:1 ratio using the nearest neighbor method based on age, sex, and BMI, using the *matchit* package.^30^ Controls were excluded if they had active gastrointestinal symptoms or diseases, and used medications affecting gastric motility or regular cannabis. Specific exclusion criteria related to BSGM (Gastric Alimetry) were BMI >35, active abdominal wounds or abrasions, fragile skin, and allergies to adhesives.

### Gastric Alimetry Protocol

Details of the Gastric Alimetry System are reported in the **Supplementary Methods** and **Fig. 1**. This system has been comprehensively validated to specifically detect gastric myoelectrical activity,^26,27^ and also registers gastric contractile activity through an increase in signal power.^31^ The standard Gastric Alimetry test protocol was followed (**Fig. 1A**). Participants were fasted for >6 hrs and avoided caffeine and nicotine prior to testing. Medications known to influence gastrointestinal motility were withheld 72 hrs prior, including opiates and prokinetics. Array placement was preceded by skin preparation (NuPrep; Weaver & Co, CO, USA). Recordings were performed for a fasting period of 30 minutes, followed by a 482 kCal meal consumed over 10 minutes, followed by a 4-hr postprandial recording. The meal consisted of Ensure (232 kcal, 250 mL; Abbott Nutrition, IL, USA) and an oatmeal energy bar (250 kcal, 5 g fat, 45 g carbohydrate, 10 g protein, 7 g fiber; Clif Bar & Company, CA, USA). Participants sat reclined in a chair and were asked to limit movement, talking, and sleeping, but were able to read, watch media, work on a mobile device, and mobilize for comfort breaks.

**Figure 1.**
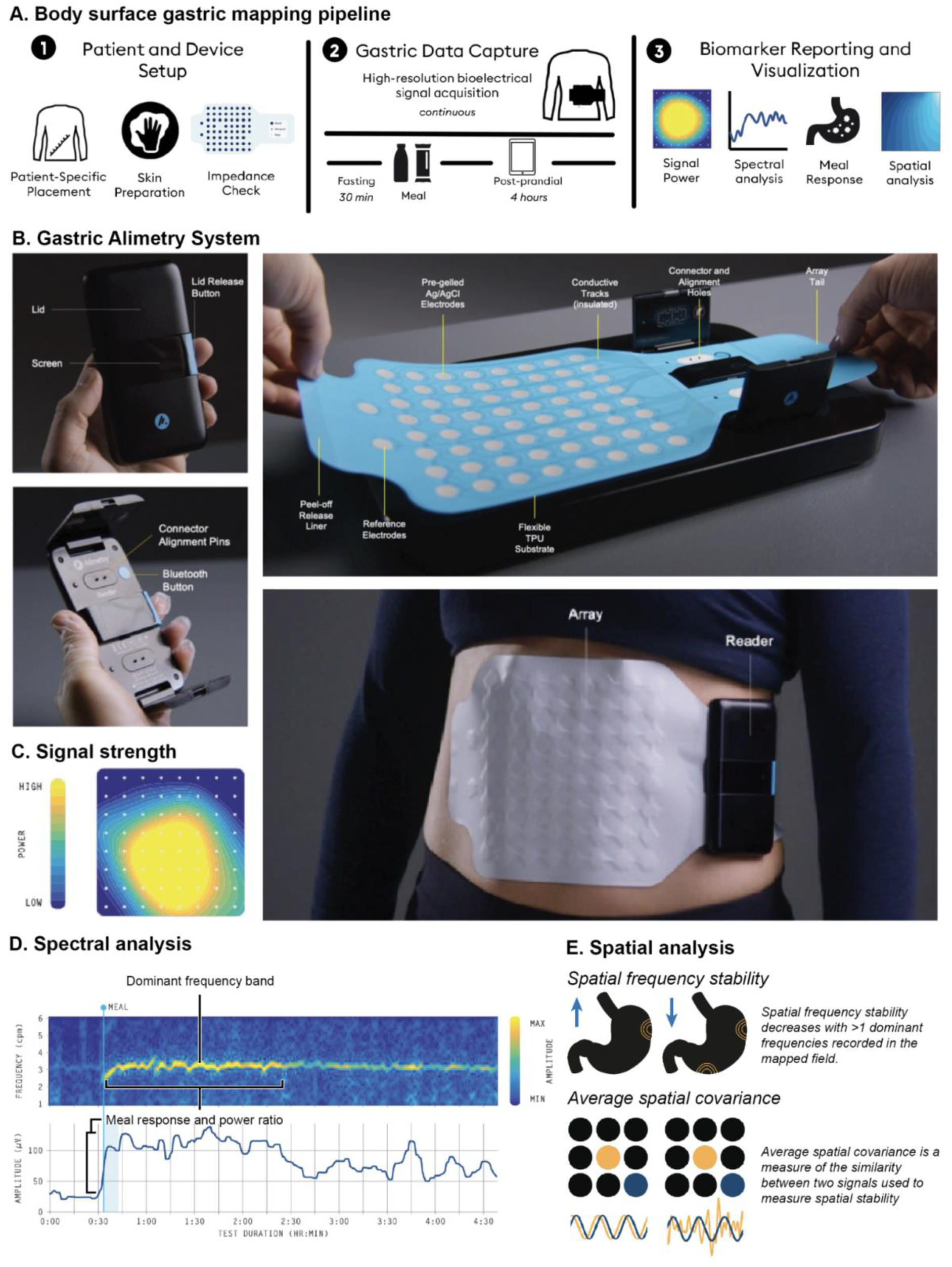
BSGM system and analysis pipeline. **A)** Outline of standardized BSGM test procedure.^27^ **B)** The Gastric Alimetry Device, shown in closed and open profiles, during setup of the high-resolution array (8×8 pre-gelled Ag/AgCl electrodes), and in position overlying the epigastrium. **C**. Signal strength is visualized on an 8×8 electrode grid corresponding to the array, with the region of high-power activity corresponding to the gastric position in the mapped field.^27^ **D**. BSGM analytics employed in this study comprised spectral metrics (meal response, power ratio, rhythm, dominant slow wave frequency; used for test classifications) and spatial metrics (measures of spatial slow wave stability; spatial frequency stability and average spatial covariance). Refer **Supplementary Methods** for further detail.

Data analytics encompassed spectral and spatial BSGM variables as described in the **Supplementary Methods**, and summarized in **Fig. 1**. As BSGM is a new test and reference ranges are still being developed, spectral data were classified by consensus panel (four expert assessors) as being normal, abnormal, or indeterminate (when no consensus reached), who were blinded to the group, with reference to established literature (refer **Supplementary Methods** and **Fig. S1**).

### Patient reported outcomes

Baseline symptom severity and quality of life were measured using the validated Patient Assessment of Upper Gastrointestinal Disorders-Symptom Severity Index (PAGI-SYM), Gastroparesis Cardinal Symptom Index (GCSI), and Patient Assessment of Upper Gastrointestinal Disorders-Quality of Life (PAGI-QOL) instruments.^32,33^ Symptoms of nausea, bloating, upper gut pain, heartburn, stomach burn, and excessive fullness were measured during testing via the validated Gastric Alimetry App at 15-minute intervals using 0-10 visual analog scales (0 indicating no symptoms; 10 indicating the worst imaginable extent of symptoms).^34^ This data was used to calculate the ‘Total Symptom Burden Score’.^34^ Early satiation was measured after the meal. Vomiting, reflux, or belching events were also recorded and assessed by frequency. Health psychology metrics included the State Trait Anxiety Inventory Short Form (STAI-SF) and Patient Health Questionnaire-2 (PHQ-2) questionnaires.^35,36^

### Sample size and statistical analysis

Assuming a 5% abnormal test rate in controls and 10% drop out, at 0.05 alpha and 90% power, a minimum of 30 NVS patients were required to detect a 25.8% difference in the abnormal test rate between NVS patients and controls.

Principal component analysis (PCA) was used to visualize data groupings based on the consensus panel classification (Python v.3.9.7 *scikit-learn* package). PCA provides a means for representing high-dimensional data in a low-dimensional space while maximally preserving the variance in the data, by finding weighted combinations of variables (i.e., principal components) that are linearly uncorrelated and can be used to transform data to obtain principal component scores. Statistical analyses were performed in R v.4.0.1 (R Foundation for Statistical Computing, Vienna, Austria). Data were expressed as mean ± SD, or SEM, or median (IQR). Student’s t-test or one-way ANOVA was used for normally distributed data and Mann-Whitney U or Kruskal–Wallis for non-normally distributed data. Post-hoc testing of ANOVA with Tukey correction was performed in subgroup analyses. Pearson correlations with Hochberg corrections for multiple testing were used to generate correlation matrices for wheel plots.^37^

## Results

Forty-three consecutively recruited NVS patients were matched to 43 controls. **Table S1** presents population demographics, past medical history, and medications. Overall 66/86 (76.7%) participants were female, median age was 33 (IQR 26 - 44), and mean BMI was 24.3 (SD 4.3). Among patients, 22 were diagnosed with gastroparesis. Patient-reported outcomes are reported in **Table S1**, and BSGM technical details in **Tables S2 and S4**. BSGM was successful in all controls and 42/43 patients, with one excluded due to excessive motion artifacts (>65% of test duration).

### Whole cohort analysis

NVS patients demonstrated multiple BSGM abnormalities compared to controls (**Fig. S2**), including impaired meal responses, characterized by attenuated average postprandial amplitudes (median 23.3 µV (IQR 18.5-31.3) vs 38.0 µV (IQR 23.9-63.2); p<0.001) and reduced fed-to-fasting power ratio (median 1.11 (IQR 0.95-1.54) vs 1.59 (IQR 1.1-2.0); p=0.02). Average dominant frequencies were normal in both groups (mean NVS 2.69 cpm (SD 0.43); controls 2.88 cpm (SD 0.36)), but patients showed greater variance (0.69 (SD 0.29) vs 0.48 (SD (0.26); p<0.001). Spatial biomarkers revealed relatively unstable myoelectrical activity in NVS, with reduced spatial frequency stability (median 13.6 (IQR 4.5-42.5) vs 49.5 (IQR 19.1-79.0); p<0.001), and reduced average spatial covariance (0.49 (SD 0.06) vs 0.53 (SD 0.06); p=0.002).

### Phenotypes within NVS cohort

Although the whole cohort analysis revealed strong differences between patients and controls, patient data were heterogeneous (**Fig. S2**). Two dominant patient subgroups were identified based on normal vs abnormal BSGM tests, which were evaluated with respect to the consensus panel classifications (**Fig. 2, S1, S3**). Among controls, 41 (95%) were classified as having normal spectral patterns (**Fig. 2A, S3A**) (including normal variants - refer **Supplementary Methods, Figs S1, S4**), one was indeterminate, and one control was classified as abnormal. Among NVS patients, 26 had normal spectral patterns (62%) (**Fig. 2B; Fig S3B**), which included 3 patients with isolated high stable frequencies or only transient instabilities (**Fig. S4**). Thirteen had abnormal spectral patterns (31%), characterized by absent meal responses, weak or inconsistent dominant frequency bands, and unstable slow-wave rhythm (**Fig. 2C; Fig. S3C**). Four NVS patients were classified as indeterminate.

**Figure 2.**
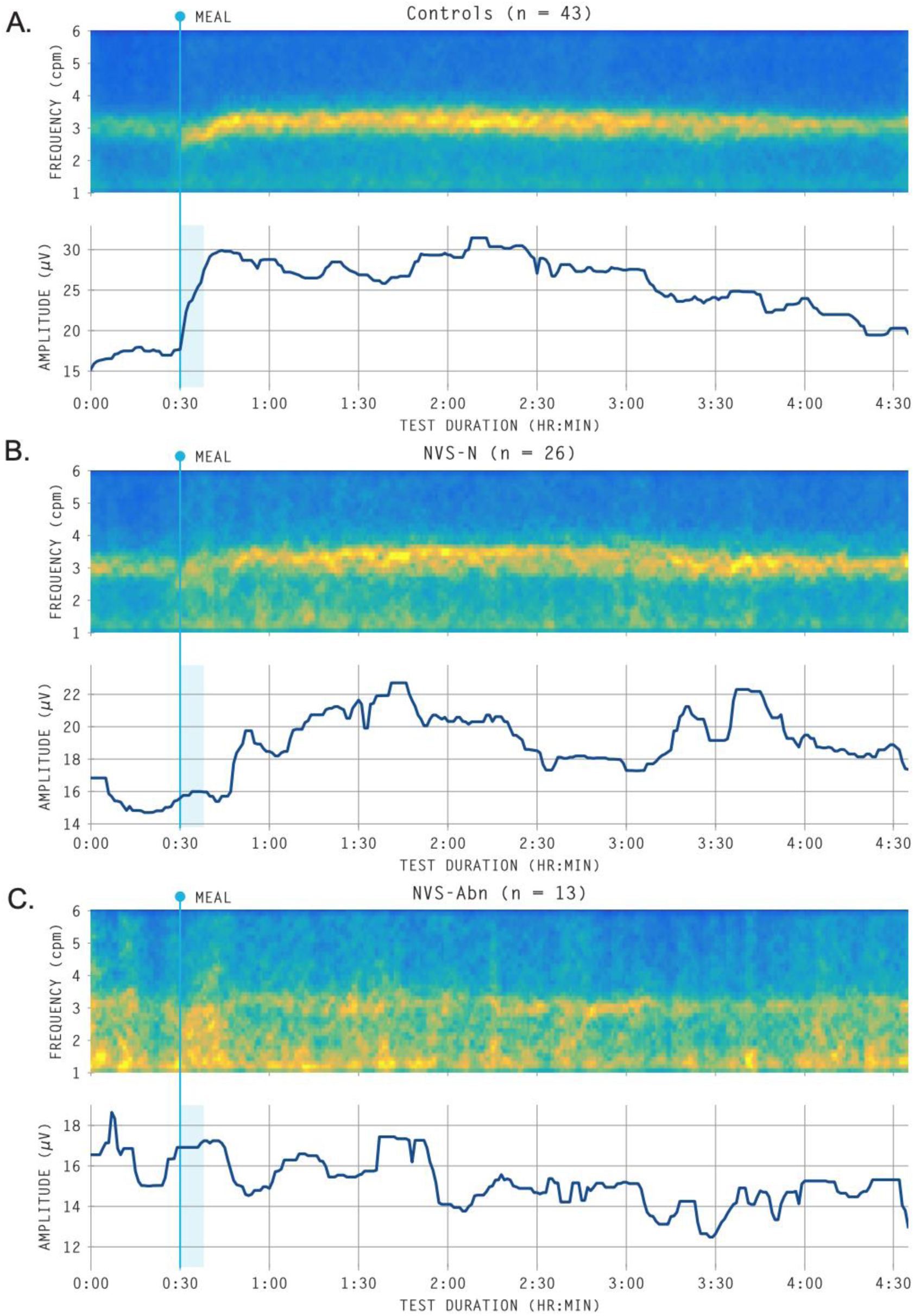
Average BSGM spectral plots (frequency-amplitude graphs), showing dominant frequencies on a scale from low power (dark blue) to high power (bright yellow), indicating gastric meal responses and rhythm, and with amplitude plots beneath, for healthy controls, NVS-N and NVS-Abn groups. Themeal time and duration is indicated by a vertical blue bar at 30 min. Normal spectral plots (**A,B**) show a clear meal response (post-prandial power increase), a consistent and sustained frequency band, and a regular gastric rhythm (**Supplementary Methods** and **Fig. S1**). **C)** Abnormal cases lacked these features. Individual subject examples from each category are provided in **Fig. S3**, and examples of normal and patient variations are provided in **Fig. S4**.

Subgroup comparisons were performed between patients with normal BSGM spectral patterns (*“NVS-N”*) vs abnormal patterns (*“NVS-Abn”*). Symptom profiles and quality of life data were comparable between subgroups (p>0.1 for all symptoms and patient-reported outcomes; **Fig. 3**, **Table S2**). However, *NVS-N* patients had substantially higher rates of clinically-diagnosed anxiety and/or depression (44.0% vs 7.7%; p=0.03), were more likely to be on anxiolytic or antidepressant / neuromodulator prescriptions (50.0% vs 15.4%; p=0.045), and had higher state anxiety scores (median 16.5; IQR 14-21 vs 13; IQR 10-16; p=0.035) (**Fig. 3**). *NVS-Abn* showed statistically different spectral and spatial BSGM metrics compared to *NVS-N* patients, with higher standard deviation of the dominant frequency (p=0.04), a lower percentage of the recording in normal gastric frequency (31% vs 76%, p=0.04), and reduced spatial frequency stability (p=0.004) (**Fig. 3**). In comparison to the control cohort, the *NVS-Abn* group had significantly impaired spectral and spatial metrics (p<0.02), whereas all metrics were comparable to controls in *NVS-N* patients (p>0.05) (**Table S2**, **Fig. 3**). Delayed gastric emptying status by subgroup is reported in **Table S4**.

**Figure 3.**
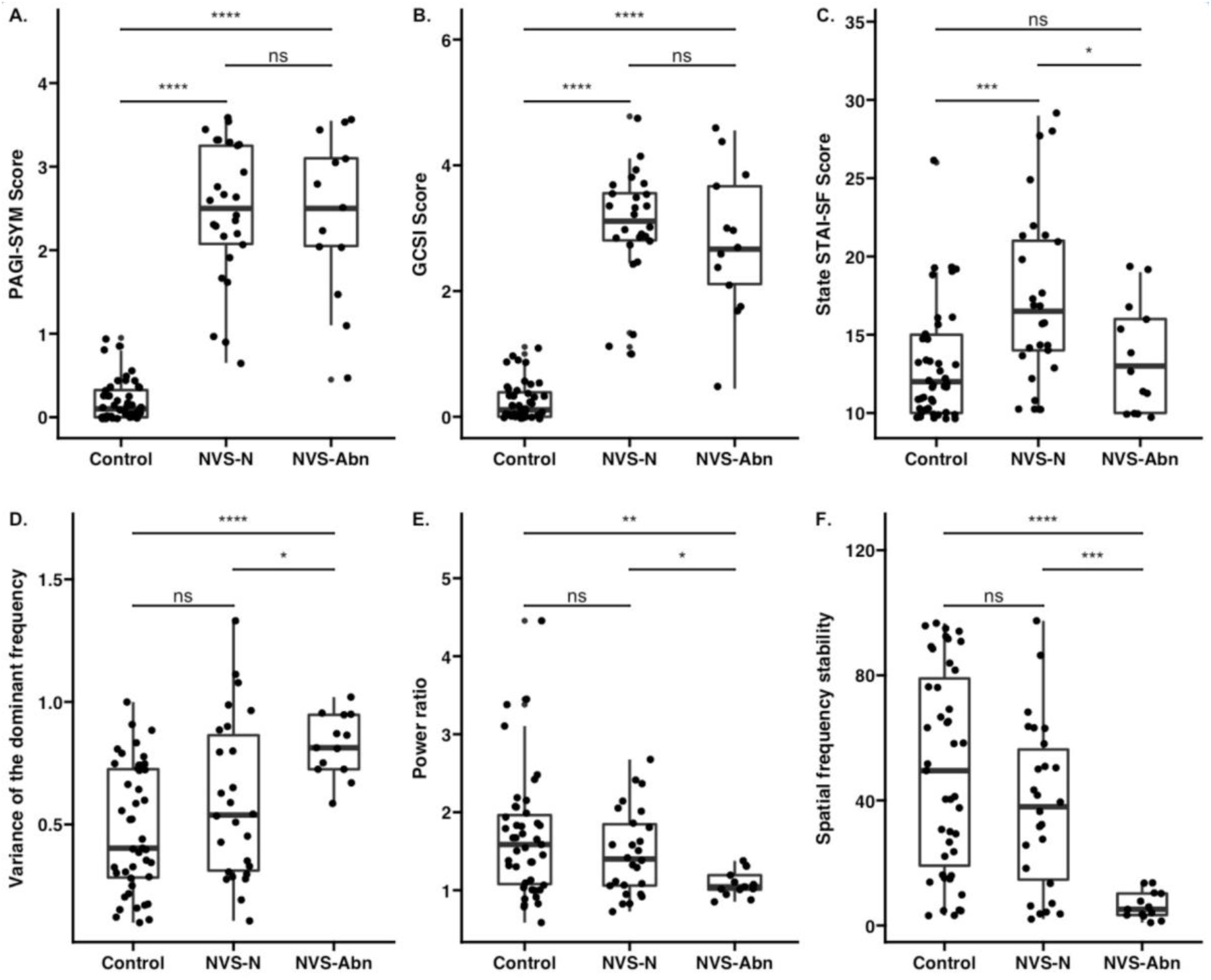
Box plots of symptoms and metrics across controls, and those that had a positive (abnormal) and negative (normal) BSGM test in the NVS cohort. PAGI-SYM, Patient Assessment of Upper Gastrointestinal Disorders–Symptom Severity Index; GCSI, Gastroparesis Cardinal Symptom Index. STAI-SF; State-Trait Anxiety Inventory Short Form. ns; p>0.05, *; p≤0.05, **; p<0.01, ***; p<0.001, ****; p<0.0001.

PCA enabled visual confirmation of group differences as a function of weighted metric combinations. A combination of BSGM, psychological questionnaires, and symptom metrics clearly separated controls, *NVS-N* and *NVS-Abn* groups (**Fig. 4**). PCA using only BSGM test metrics separated *NVS-Abn* patients from *NVS-N* patients and controls, whereas conducting PCA using only psychological questionnaires and symptom metrics separated patients and controls, but did not separate *NVS-N* and *NVS-Abn* patients (**Figs. S5 and S6**).

**Figure 4.**
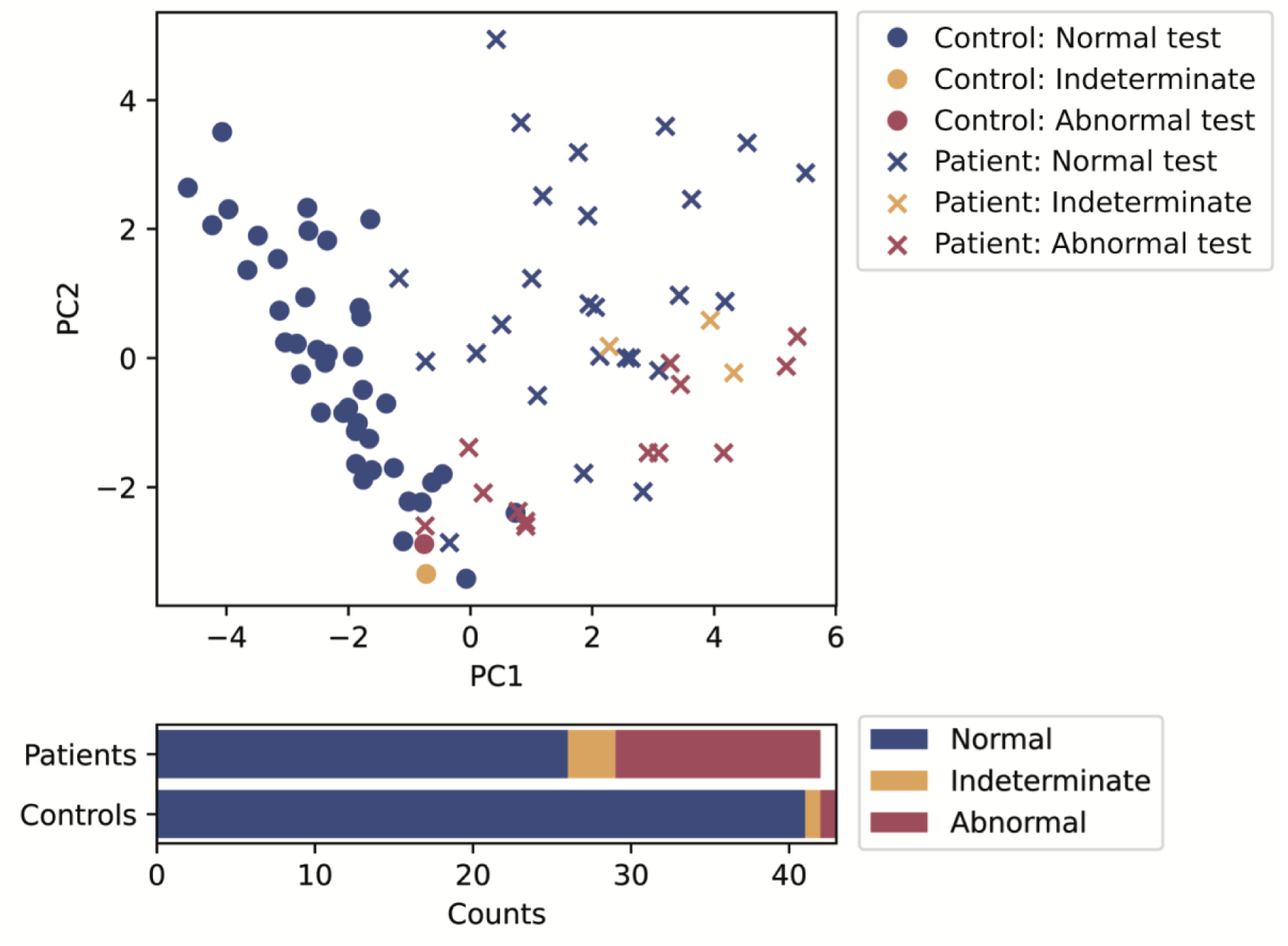
**Top**: Principal component analysis of healthy controls and NVS patients utilizing body surface gastric mapping metrics, symptom scores, and health psychology data. Below: case classifications.

#### Symptom Correlations

Symptom correlations were performed between the *NVS-N*and *NVS-Abn* subgroups and controls. In the *NVS-N* group, anxiety and depression metrics correlated with symptom severity (r>35; p<0.05; **Fig. 3** and **Fig. 5A**), whereas BSGM metrics did not correlate (p>0.4). In the *NVS-Abn* group, BSGM metrics correlated with the severity of symptoms (including nausea, pain, excessive fullness, early satiety, bloating, heartburn and GCSI score; all r>0.35, p<0.05), whereas anxiety and depression metrics did not correlate (p>0.1) (**Fig. 3** and **Fig. 5B**). In the *NVS-Abn* group, the % duration in the normal frequency range was strongly correlated with mean GCSI (r=-0.58; p<0.001) and PAGI-SYM (r=-0.59; p<0.001),

**Figure 5:**
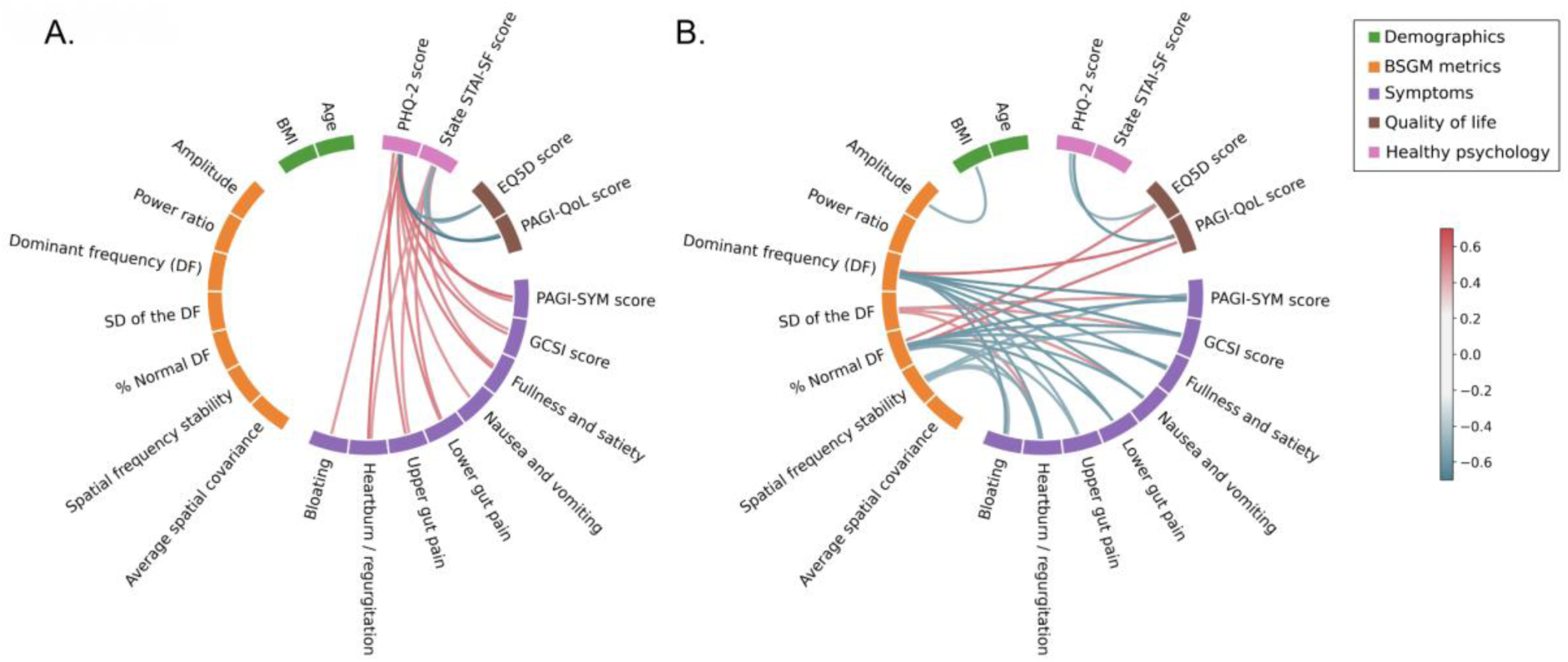
Wheel plot showing correlations between demographics, BSGM metrics, symptoms as measured by the PAGI-SYM subscales, quality of life and health psychology metrics in NSV groups. Correlations between quality of life metrics and symptoms are not reported, and only statistically significant correlations between categories are shown (after Hochberg correction). A: *NVS-N* cohort and controls; B: *NVS-Abn* cohort and controls. BSGM, body surface gastric mapping, SD; standard deviation, DF; dominant frequency, GCSI; gastroparesis cardinal symptom index, PAGI-SYM, Patient Assessment of Upper Gastrointestinal Disorders–Symptom Severity Index; EQ5D; EuroQoL 5, EuroQoL 5D, PAGI-QoL; Patient Assessment of Upper GastroIntestinal Disorders-Quality of Life, STAI-SF; State-Trait Anxiety Inventory Short Form, PHQ-2; Patient Health Questionnaire-2.

### Safety

No participants experienced a serious adverse event. Mild transient side effects included rash (n=12) and itch (n=8), with one patient requiring topical therapy. The test was well tolerated overall, evidenced by patients being highly likely to recommend the test to other patients when asked on a scale of 0 to 10 (median 9; IQR 8-10).

## Discussion

This study introduced a new medical device and test for non-invasively characterizing gastric myoelectrical activity using BSGM, and applied it to NVS. Multiple abnormalities were revealed including weak, irregular and unstable gastric myoelectrical activity and impaired meal responses. These abnormalities were confined to a subgroup (31%), with 62% of patients demonstrating normal tests, as confirmed by group-wise statistics, PCA, and symptom correlations via linear regression analysis. These data indicate that NVS likely comprises distinct underlying phenotypes that can be differentiated by BSGM.

Gastric neuromuscular pathology and dysregulation of brain-gut interaction have emerged as dominant putative mechanisms of NVS.^15,19,38^ Our findings are consistent with both mechanisms, but suggest that each may be responsible for a separate phenotype; a distinction not previously possible because of overlapping symptom profiles. Although ICC or immune cell counts were not measured in this non-invasive study, it is likely that the BSGM abnormalities observed are the electrophysiological correlate of underlying neuromuscular disease. Multiple studies have identified ICC network depletion as a pathological hallmark of both gastroparesis, and more recently CNVS, associated with a macrophage-driven immune dysregulation.^11,16–19^ ICC loss leads to slow-wave irregularities and contractile dysfunction,^16,17,20^ which is consistent with our data showing weak, irregular and unstable gastric myoelectrical activity and impaired meal responses. Benchmark studies have shown that ICC depletion is restricted to ∼50% of NVS patients, with the remainder showing normal ICC counts,^16,19^ approximating our two subgroups. As evaluating myenteric ICC currently requires highly-invasive full-thickness gastric biopsies, the development of BSGM as a non-invasive tool for the routine clinical assessment of gastric motility could represent a valuable clinical advance.

The subgroup with normal BSGM tests were significantly more likely to have a clinical diagnosis of anxiety and depression and had higher anxiety state scores, consistent with dysregulated brain-gut interaction.^15^ Brain-gut symptom genesis is well established, including in NVS,^39^ such that it has been proposed to rename functional gastrointestinal disorders to disorders of brain-gut interaction.^15^ However, neither descriptor appears adequate in this context, because of the subgroup with organic neuromuscular disease.^11,16^ In addition, approximately half of the patients with normal BSGM tests in our study showed no psychological comorbidities, and there are other mechanisms (e.g. diabetic neuropathy) to account for the heterogeneity of NVS.^40^ With further validation, BSGM could support the evolution of NVS nomenclature toward more specific disease subtypes, which in turn should improve clinical decision making (e.g. selection of targeted treatments) and allow more homogenous allocation into research trials.

Patients with NVS have previously been divided into those with gastroparesis or CNVS based on the presence or absence of delayed gastric emptying. However, this division is increasingly tenuous, given that epidemiology, symptom profiles, and quality of life impacts are comparable, and that gastric emptying may not be a primary disease mechanism.^6,7,11,41^ Moreover, Pasricha et al recently demonstrated that gastric emptying rates are labile over time, with >40% of patients switching disease categories within one year. In view of this poor specificity, NVS patients were considered here as a common group. To determine how BSGM and gastric emptying testing compare, it will be necessary to perform the two tests simultaneously.

Alvarez introduced EGG 100 years ago in a seminal paper in JAMA,^42^ and multiple EGG studies have demonstrated aberrant gastric myoelectrical activity in >50% of NVS patients, predominantly based on frequency endpoints.^13^ However, clinical adoption of EGG has not followed because it has generally been considered unreliable for investigating individual patients or guiding management, partly related to difficulties in interpreting the signals.^43^ BSGM by Gastric Alimetry represents a critical advance over EGG, enabled by multiple technical improvements including modern bioamplifiers, HR flexible electronics, validated signal processing algorithms for automated noise reduction, and a patient-specific electrode positioning guide to address anatomical variation.^22,27,44^ Four hour standard post-prandial recordings were also introduced, because we found that meal responses in controls show variable time courses that could confound matched comparisons.^27^ In addition, new endpoint criteria were established, including a number of novel biomarkers such as spatial parameters, after previously determining that frequency alone is insufficient to characterize gastric abnormalities.^17,22,27,45^ Taken together, these improvements yielded sufficient discriminative power to clearly distinguish a subgroup of patients with gastric dysfunction amongst NVS patients by multiple statistical methods and at the individual subject level.

Our results with a medical device improve upon earlier reports using BSGM research prototypes.^24,25^ Gharibans et al studied a mixed cohort including functional dyspepsia, and reported that abnormal slow-wave direction correlated with symptoms.^24^ For NVS, we focused primarily on slow-wave stability and meal response metrics, because our previous invasive HR mapping studies documented slow-wave irregularities in association with ICC depletion.^16,17^ However, it would now be valuable to comprehensively study functional dyspepsia for comparison. Somarajan et al applied a prototype BSGM system in a cohort of children with NVS, finding aberrant frequencies and spatial patterns,^25^ indicating that Gastric Alimetry will also be an applicable non-invasive tool in pediatric populations.^46^

Several limitations are acknowledged. Although specific criteria were employed, using a consensus panel classification introduces subjectivity that could limit reproducibility. BSGM reference ranges are therefore being established to promote standardized reporting. Classifications were conservative, being based primarily on meal responses and gastric rhythm, and patients with borderline tests (e.g. isolated high stable frequencies and transient abnormalities) were currently classed as negative. As more data becomes available, it will be possible to refine the classification scheme, and together with emerging spatial metrics,^26,47^ this should increase discriminative power.

In summary, this study introduces a novel medical device for BSGM and demonstrates its utility in a study of NVS patients who share common symptom profiles but reveal distinct phenotypes. These findings could improve clinical management of NVS patients by separating those with gastric dysfunction from those with gut-brain dysregulation or other aetiologies.

## Data Availability

Data produced in the present study are available upon reasonable request from the corresponding author.

## Conflicts of Interest

GOG, PD and AG hold grants and intellectual property in the field of gastrointestinal electrophysiology. GOG and AG are founders and Directors of Alimetry Ltd. GOG is Director in The Insides Company. SC, SW, CD, GS, PD, JW and CNA are members of Alimetry Ltd. PD is a Director in FlexiMap Ltd. The remaining authors have no conflicts of interest to declare.

## Funding

This work was supported by the Health Research Council of New Zealand, Royal Society Te Apārangi, and the John Mitchell Crouch Fellowship from the Royal Australasian College of Surgeons.

## Acknowledgements

We thank Gen Johnson, India Wallace, Sonia Binden, Lynn Wilsack, and Renata Rehak for their outstanding assistance with participant recruitment, and to all of our study participants for enabling this work.

## Supplementary Appendix

### Methods

#### Gastric Alimetry® System

Gastric Alimetry® is a novel medical device custom-designed for BSGM and detailed here in full for the first time. The device consists of an HR Array, wearable Reader, Dock, iPadOS App for setup and symptom logging, and cloud-based analytics and reporting platform (**Fig. 1A**). Key design considerations for each component are detailed below.

- *Array*. The Gastric Alimetry Array™ includes 66 pre-gelled Ag/AgCl electrodes (8×8 grid +2 reference; inter-electrode spacing 20 mm), covering an area of 21×16 cm (196 cm^2^) (**Fig. 1B**). This Array™ specification was designed to overlie the majority of the stomach’s area in >95% of cases, which is important because gastric position is highly variable and the weak gastric signal strength diminishes exponentially from source.^48–50^. Each Array is single-use, being screen printed using conductive inks on a single, flexible, thermoplastic polyurethane (TPU), with an overlying peel-and-stick adhesive layer that enables rapid setup and removal.^27^
- *Reader.* The Alimetry Reader™ incorporates custom-designed electronics specifically tuned for gastric electrophysiology (**Fig. 1B**). Signals are acquired at 250 Hz, then amplified and digitized by low-noise programmable gain amplifiers, with each input compared against a common reference electrode to provide unipolar recordings, while movement artifacts are registered by an onboard accelerometer. The Reader attaches to the Array using a custom board-to-board connector design that eliminates all cabling to enable unimpeded wearability and facilitate ease of cleaning.
- *Dock.* The Alimetry Dock™ is used for charging and storage of the Reader, and accurate alignment of the Array during setup (**Fig. 1B**).
- *App*. The Gastric Alimetry App™ runs on an iPad mini (Apple, Cupertino, CA), and is used for device setup, data transfers, and to capture patient-reported symptom data during testing. Guided setup in the App includes an Array positioning step that tailors placement per individual patient biometrics, to further enhance accurate positioning over the stomach.^27^ Patients log symptoms every 15 minutes via a digital interface employing pictograms (**Fig. 1A**), which has been validated to enable reliable capture of patient symptom data in association with a standard meal with excellent compliance.^34^ This system therefore enables precise temporal correlations between patient symptom profiles with electrophysiological data.
- *Cloud / Portal.* Test data is transmitted to a HIPAA-compliant cloud server at the conclusion of each test. A proprietary algorithm automatically filters and analyzes raw myoelectrical signals to generate a report, including key metrics and data visualizations (detailed below), which are accessible via a secure online portal. Filtering methods are based on a previously validated scheme by Gharibans et al, accounting for accelerometer data.^44^

#### Spatial and spectral data analytics

##### Spectral analysis

Spectral metrics represent the bioelectrical slow-waves that coordinate gastric motility, as well as gastric contractile activity registered through an increase in signal power, and are visualized using spectral plots (**Fig. 1D**).^31,51^ They are comparable to traditional EGG metrics, but are substantially more accurate due to the higher signal-to-noise ratio (SNR) of BSGM, which is enabled by a greater coverage over the stomach’s area, reliable positioning over the stomach to account for anatomical biodiversity, summation of spectral data over a large number of active sensors, use of modern bioamplifiers, and validated signal processing techniques to account for noise including competing biological sources.^22,24,27,44,48^ These metrics encompass mean amplitude (microvolts; μV), dominant frequency (cycles per minute; cpm), standard deviation (SD) of the dominant frequency, and the power ratio (the ratio of the mean amplitude in the fed vs fasted phase).^52^ In this study, these variables were visualized for the entire test period, but calculated in statistical analyses based on the first 2 hr postprandially, when gastric activity is typically most active and SNR is highest.^27^

##### Spatial analysis

These metrics encompass spatial descriptions of gastric electrophysiological activity, providing increased sensitivity by detecting abnormal gastric activation patterns.^16,17,20,22,24,45^ The spatial metrics assessed in this study were devised to measure wave propagation stability, which was previously shown to be abnormal in subsets of NVS patients in serosal HR mapping studies.^16,17^ These spatial metrics included the ‘spatial frequency stability’ (defined as the percentage of the recording during which there is a single dominant frequency across the mapped field), ‘average spatial covariance’ (defined as the average absolute value of the covariance between pairs of electrodes computed over the course of a recording).

#### BSGM test interpretation framework

Test quality was assessed by the consensus panel on a scale of 1 to 4, incorporating the following features according to the Gastric Alimetry labeling.

a. successful stomach localization (>7 electrodes localized within mapped fields)^27^
b. impedance (<200 kΩ optimal; <500 kΩ acceptable; 500+ kΩ poor)
c. proportion of artifacts (<20% optimal; <50% acceptable; >50% interpret with caution)

- Tests of score 1 were excluded, tests of score 2 were included but with caution advised in classification as normal or abnormal, and tests of 3-4 were included with confidence.

Reference ranges and spatial metrics for BSGM are still under development, and have not yet achieved regulatory approval for the Gastric Alimetry device. Classifications of individual subjects as normal, abnormal or indeterminate tests were therefore undertaken by the consensus panel exclusively according to spectral analyses in this study. The following 3 features were employed, according to the Gastric Alimetry labeling and existing gastric electrophysiology literature:^20,27,52,53^

a. presence of a meal response (power ratio >1.4 compared to a baseline period)
b. presence of a clear and consistent dominant frequency band within the spectral plot
c. presence of a normal rhythm (stability typically within a range of 2.5 - 3.6 cpm)

- Isolated stable high or low frequencies were not judged abnormal in this study.^16,17,45^

The application of this process is summarized in **Supplementary Figure S1**.

#### Supplementary statistical methods

Where there was missing data for components of PCA analysis, the average value of the cohort was taken and substituted (overall there were fewer than n = 4 missing data points).

**Table S1:**
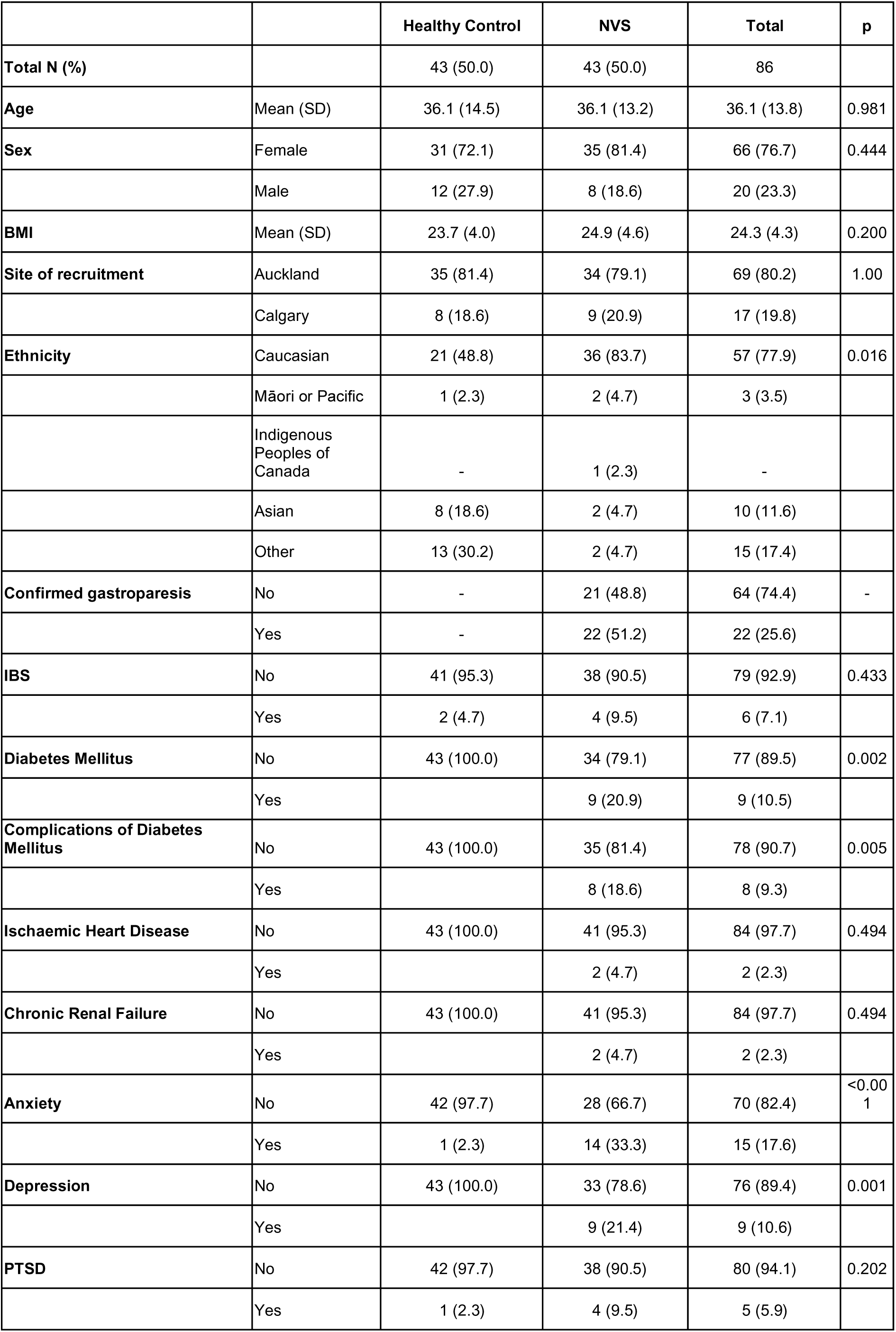
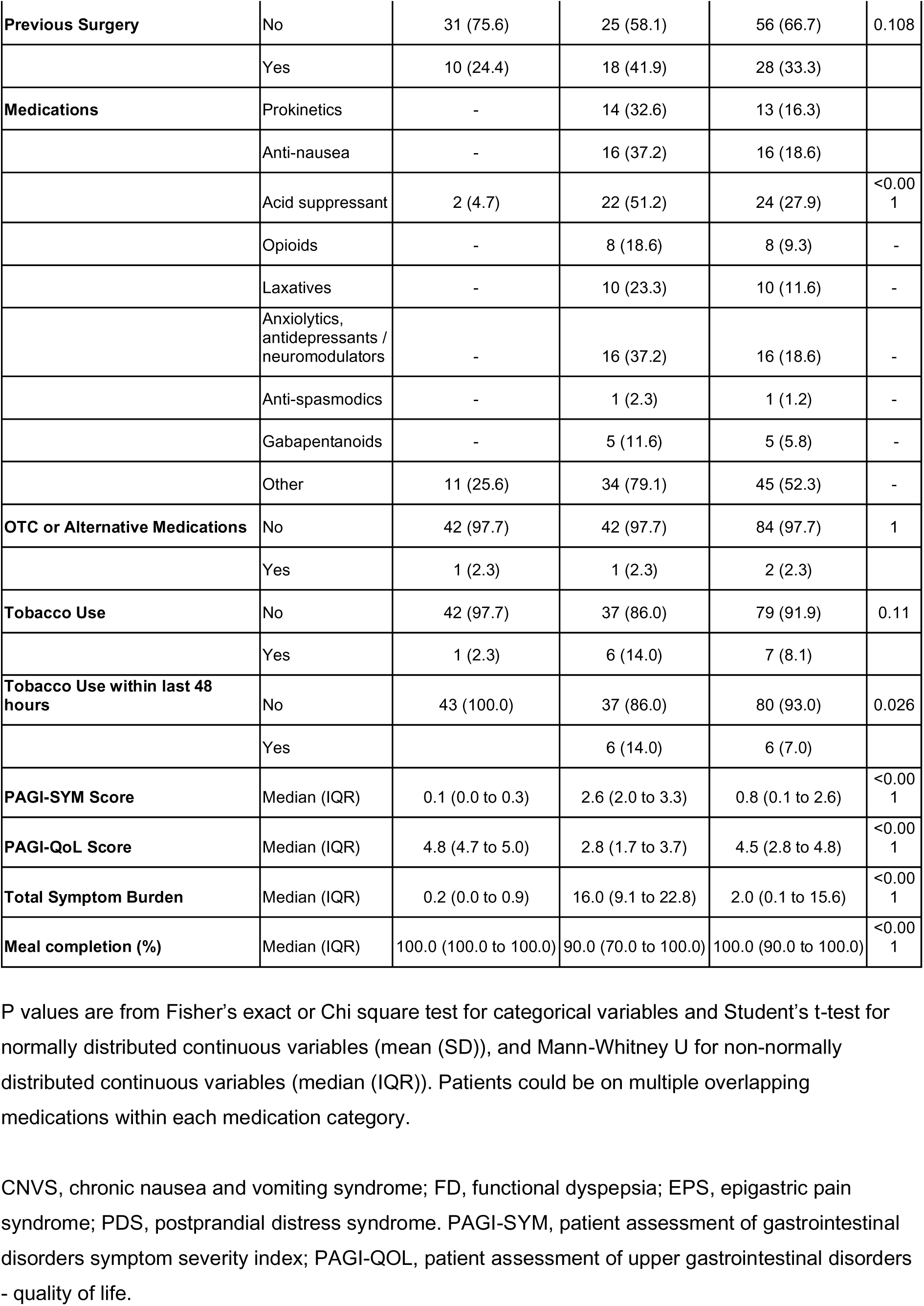
Cohort demographics

**Table S2:** Symptoms, quality of life, spectral and spatial data between controls and phenotyped NVS patients

|  | Control<br>(n = 43) | Gastric<br>dysfunction<br>(n = 13) | Patient with<br>normal BSGM<br>(n = 26) | P (gastric<br>dysfunction vs<br>patients with<br>normal BSGM) | P (healthy<br>controls vs<br>patient with<br>normal BSGM) | P (healthy<br>controls vs<br>gastric<br>dysfunction) |
| --- | --- | --- | --- | --- | --- | --- |
| **PAGI-SYM Score** | 0.1 (0.0 to 0.3) | 2.5 (2.0 to 3.1) | 2.5 (2.1 to 3.2) | 0.97 | <0.001 | <0.001 |
| **Total symptom burden** | 0.2 (0.0 to 0.9) | 11.7 (3.4 to 23.1) | 16.2 (11.0 to 27.6) | 0.62 | <0.001 | <0.001 |
| **GCSI** | 0.1 (0.0 to 0.4) | 2.7 (2.1 to 3.7) | 3.1 (2.8 to 3.6) | 0.49 | <0.001 | <0.001 |
| **EQ-5D** | 1.0 (1.0 to 1.0) | 0.8 (0.8 to 0.9) | 0.8 (0.7 to 0.9) | 0.1 | <0.001 | 0.006 |
| **PAGI-QoL Score** | 4.8 (4.7 to 5.0) | 3.5 (1.8 to 3.7) | 2.4 (1.6 to 3.4) | 0.27 | <0.001 | <0.001 |
| **Meal completion (%)** | 100 (100 to 100) | 100 (70 to 100) | 95.0 (70.0 to 100.0) | 1.00 | 0.01 | 0.06 |
| **Amplitude** | 38.0 (23.9 to 63.2) | 20.6 (18.1 to 24.8) | 26.7 (21.4 to 36.1) | 0.004 | 0.11 | 0.25 |
| **Variance of the dominant frequency** | 0.40 (0.3 to 0.7) | 0.81 (0.73 to 0.95) | 0.54 (0.31 to 0.86) | <0.001 | 0.18 | 0.04 |
| **Percentage duration of recording in the normal gastric frequency (2.5 – 3.6 cpm)** | 81.6 (63.1 to 92.0) | 31.3 (27.3 to 51.7) | 76.4 (23.5 to 89.4) | <0.001 | 0.07 | 0.04 |
| **Power ratio** | 1.59 (1.1 to 2.0) | 1.04 (1.0 to 1.2) | 1.4 (1.1 to 1.8) | 0.014 | 0.43 | 0.18 |
| **Spatial frequency stability** | 49.5 (19.1 to 79.0) | 5.17 (3.3 to 10.2) | 38.0 (14.7 to 56.3) | <0.001 | 0.23 | 0.004 |
| **Average spatial covariance** | 0.52 (0.49 to 0.54) | 0.47 (0.46 to 0.51) | 0.50 (0.47 to 0.53) | 0.003 | 0.14 | 0.7 |

\*All metrics in this study were measured during the first 2-hours of postprandial recording where the SNR is typically highest (refer **Supplementary Methods**). All data are reported as median (IQR). BSGM, body surface gastric mapping. Post-hoc Tukey test adjusted p values are reported.

**Table S3:** Artifact and impedance measures

|  |  | Control | Patient | Total | p |
| --- | --- | --- | --- | --- | --- |
| Total N (%) |  | 43 (50.0) | 43 (50.0) | 86 |  |
| Marked artifact | Median (IQR) | 8.1 (4.0 to 11.4) | 15.5 (7.6 to 24.6) | 9.7 (5.1 to 18.2) | <0.001 |
| Impedance | Mean (SD) | 95.6 (55.9 to 134.1) | 122.5 (83.9 to 182.7) | 112.3 (63.3 to 154.1) | 0.028 |

**Table S4:** Delayed gastric emptying status by phenotyped subgroup

|  |  | Control | NVS-N | NVS-Abn | p |
| --- | --- | --- | --- | --- | --- |
| Confirmed gastroparesis | No | 43 (100.0) | 9 (34.6) | 10 (76.9) | <0.001 |
|  | Yes | 0 (0.0) | 17 (65.4) | 3 (23.1) |  |

## Supplementary Figures

**Figure S1.**
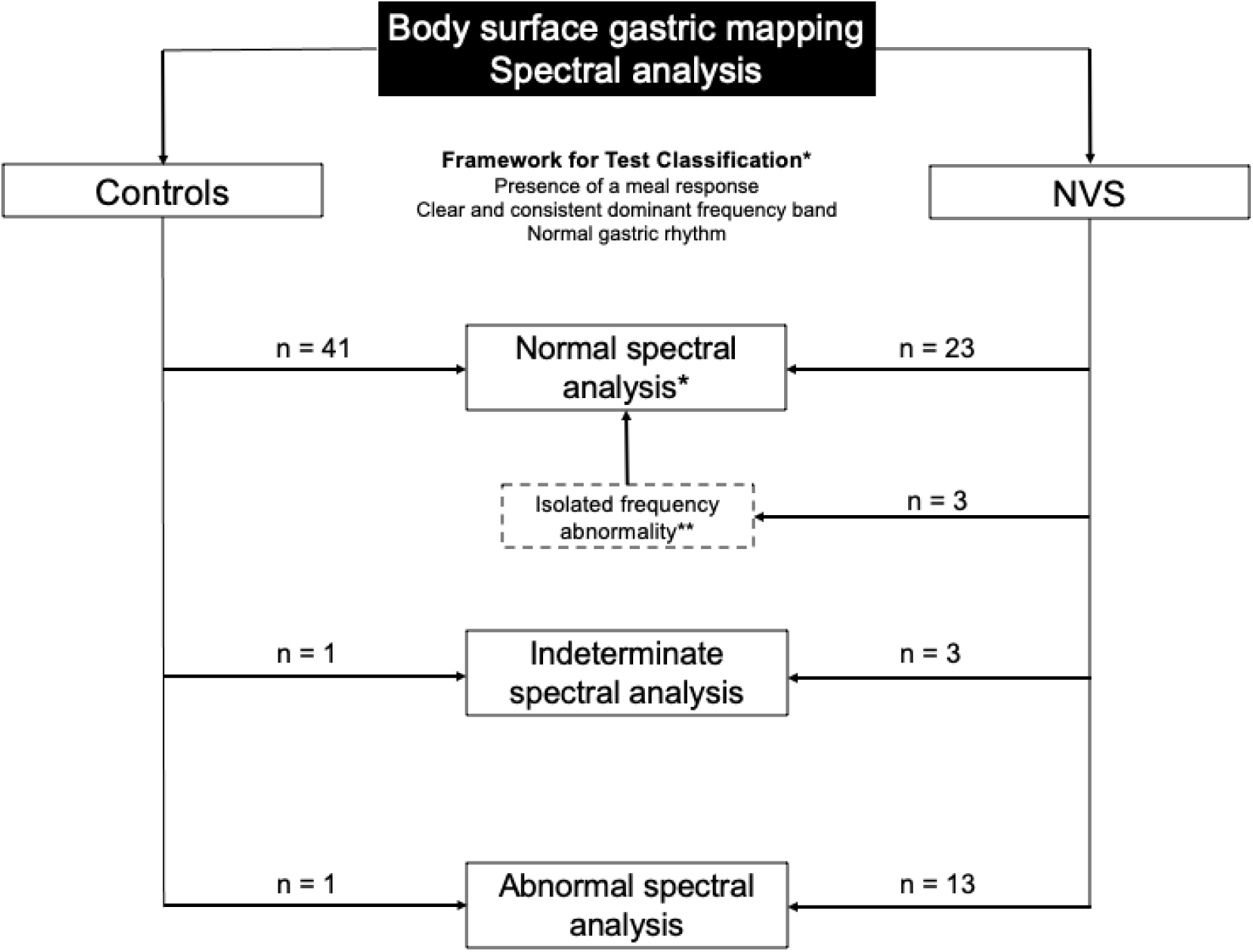
Classification scheme employed for controls and NVS patients. Among the participants that had a normal spectral analysis, there were several variants of normal characterized by either: high fasting baseline power (n = 4), limited meal response (n = 2), delayed-onset meal response (n = 5), or transient spectral abnormalities (n = 2). One NVS patient was excluded due to inadequate test quality. Data on variants and the excluded subject are provided in **Fig. S4**.

**Figure S2.**
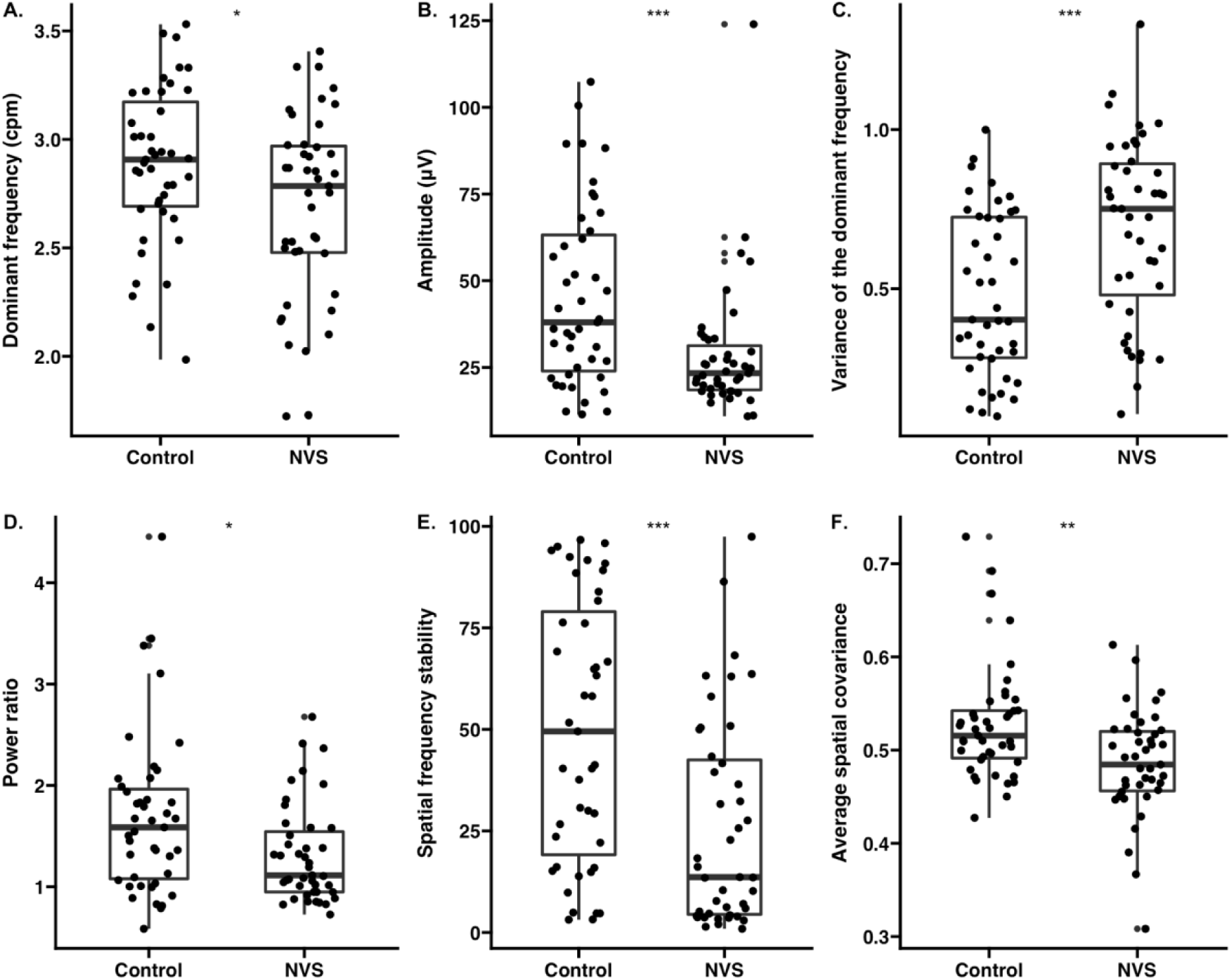
Box plots of symptoms and BSGM metrics in controls and NVS patient cohorts. cpm: cycles per minute; NVS: nausea vomiting syndrome.

**Figure S3.**
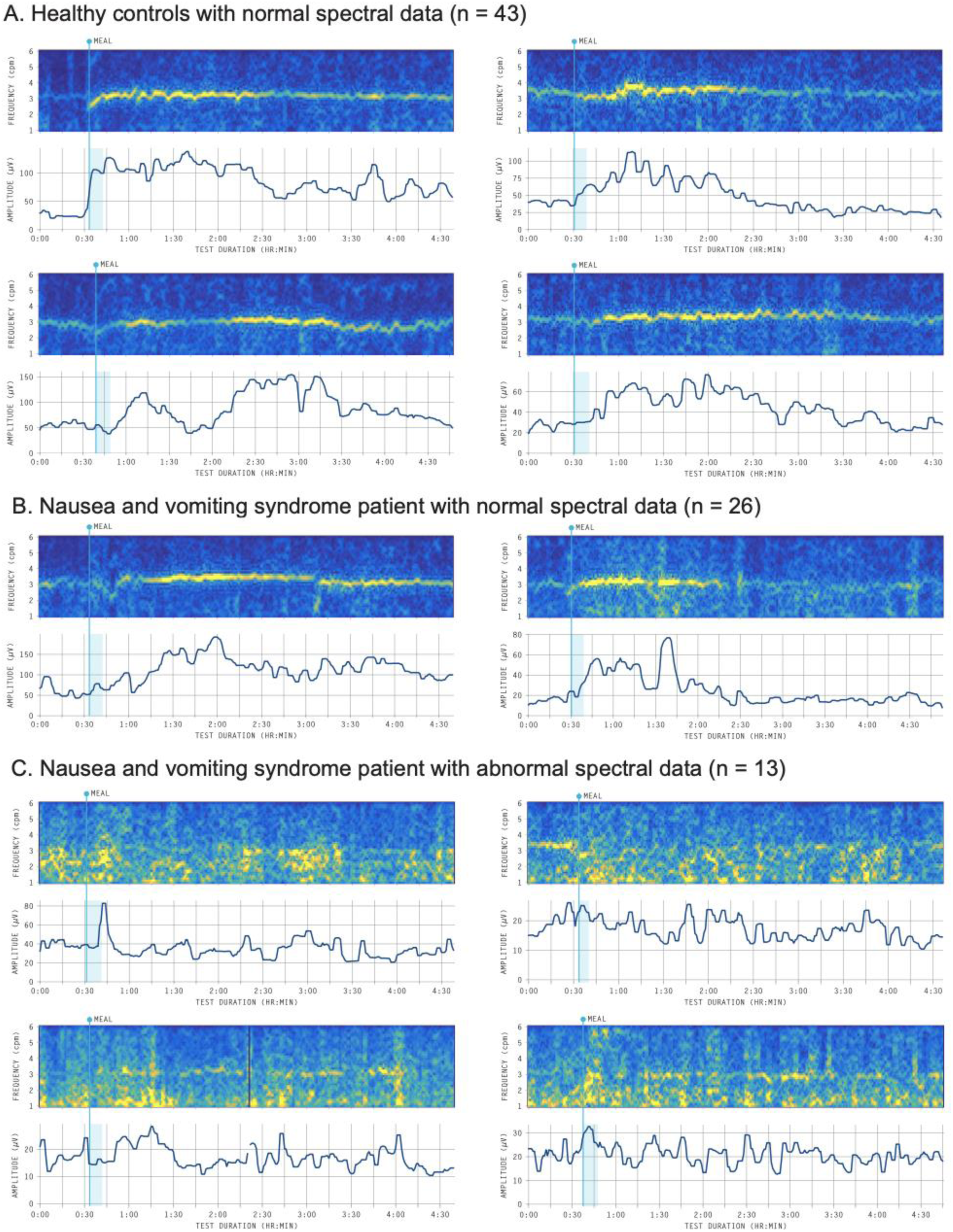
BSGM spectral plots (frequency-amplitude graphs), showing dominant frequencies on a scale from low power (dark blue) to high power (bright yellow), indicating gastric meal responses and rhythm, and with amplitude plots beneath. The meal time and duration is indicated by a vertical blue bar. Normal spectral plots (**A,B**) show a clear meal response (post-prandial power increase), a consistent and sustained frequency band, and a regular gastric rhythm (**Supplementary Methods** and **Fig. S1**). **C)** Abnormal cases lack these features. Examples of normal variants are provided in **Fig. S4**.

**Figure S4.**
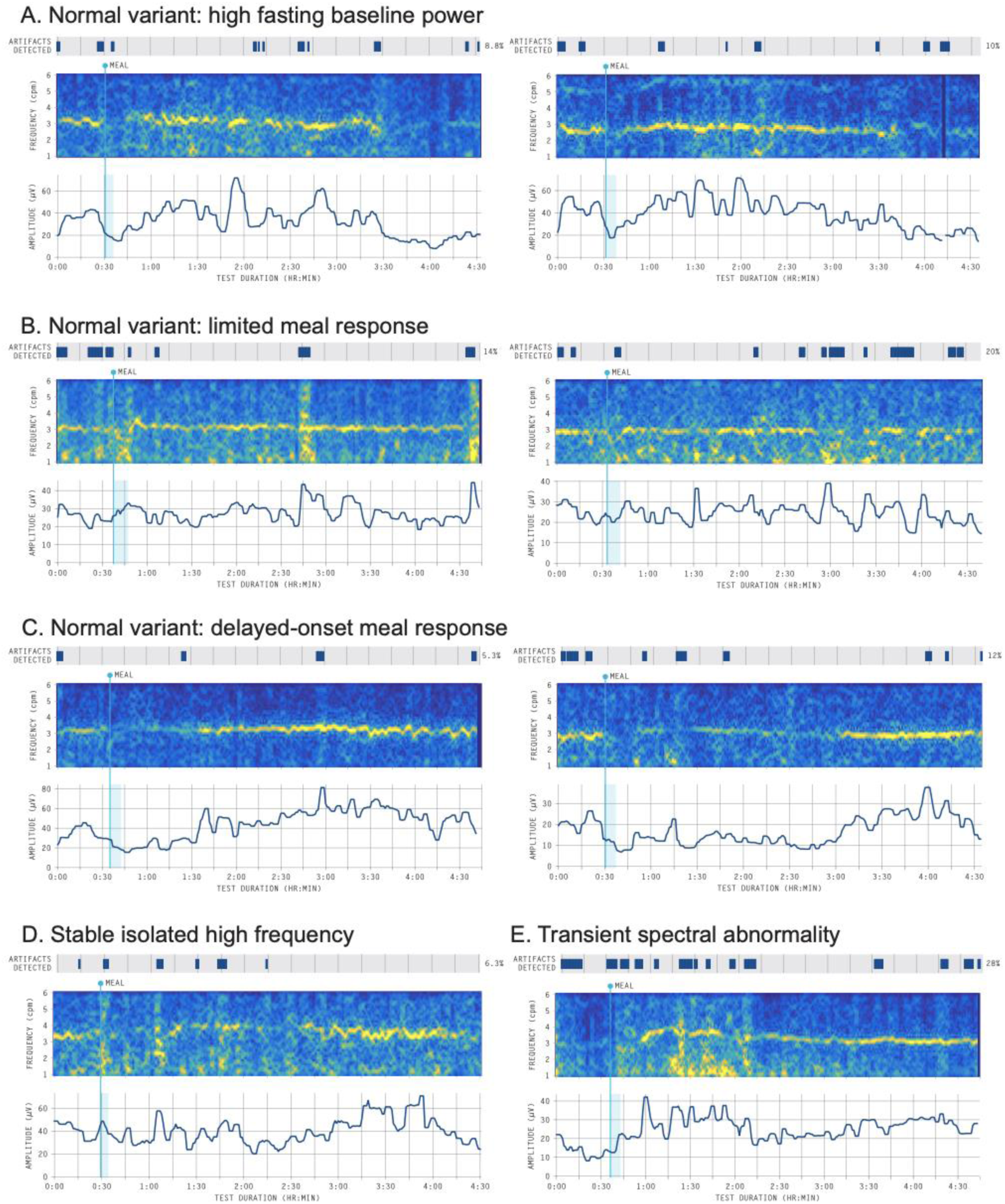
BSGM spectral plots (frequency-amplitude graphs) showing atypical data. Three variations of normal were observed in control data, as follows: **A)** High fasting baseline power (n=4); **B)** Limited meal response (n=2; no post-prandial power increase observed, but strong dominant frequency band with normal rhythm); **C)** Delayed-onset meal response (n=5). In addition, two types of abnormalities were seen in patients that did not reach threshold for an abnormal classification in this study, but were not observed in controls and may represent pathological variations: **D)** Isolated stable high frequencies (n=3; range 3.8 - 4.0 cpm); **E)** Transient spectral abnormalities (n=2).

**Figure S5.**
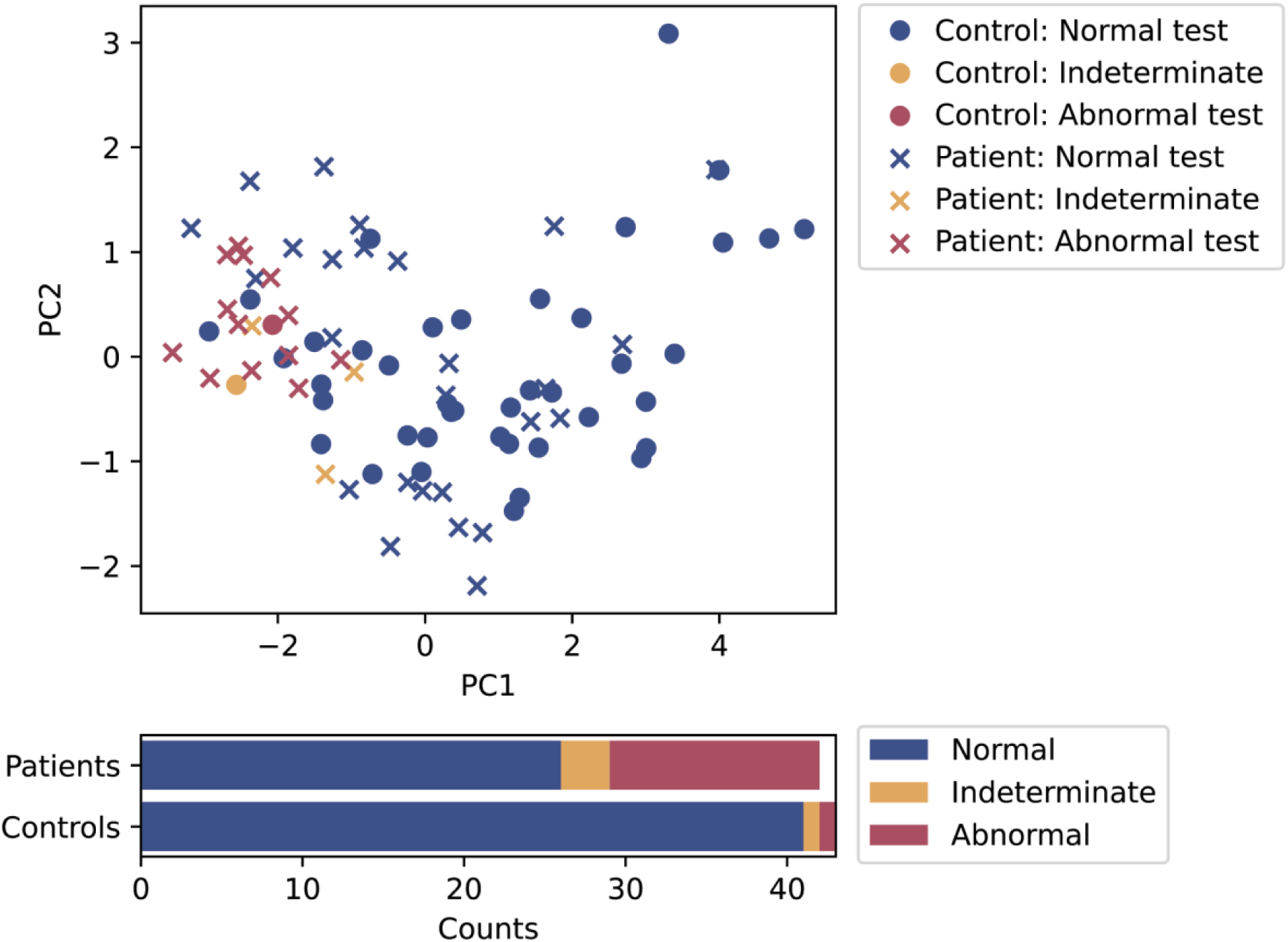
Principal component analysis including only body surface gastric metrics.

**Figure S6.**
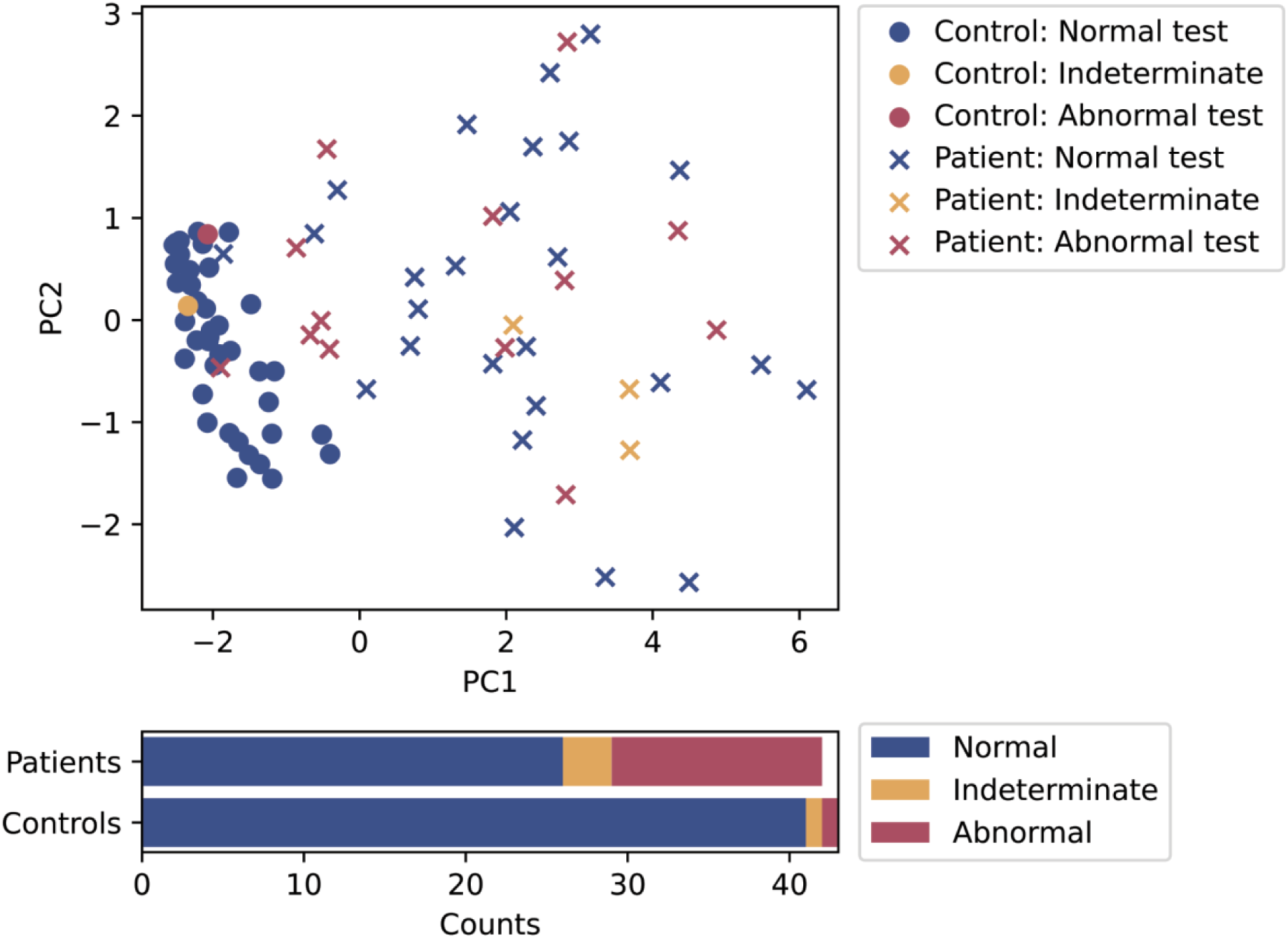
Principal component analysis including only symptoms and psychological data.

## Notes

### Author Declarations

This was a case-control study conducted in Auckland, New Zealand, and Calgary, Canada. Ethical approvals were given by The Auckland Health Research Ethics Committee (AHREC; AH1130) and Conjoint Health Research Ethics Board (CHREB; REB19-1925). All patients provided informed consent.

